# Impact of a nighttime curfew on overnight mobility

**DOI:** 10.1101/2021.04.04.21254906

**Authors:** Amir Ghasemi, Nick Daneman, Isha Berry, Sarah A. Buchan, Jean-Paul Soucy, Shelby Sturrock, Kevin A. Brown

## Abstract

**Background:** Among non-pharmaceutical interventions, individual movement restrictions have been among the most impactful methods for controlling COVID-19 case growth. While nighttime curfews to control COVID-19 case growth have been implemented in certain regions and cities, few studies have examined their impacts on mobility or COVID-19 incidence. In the second wave of COVID-19, Canada’s two largest and adjacent provinces implemented lockdown restrictions with (Quebec) and without (Ontario) a nighttime curfew, providing a natural experiment to study the association between curfews and mobility.

**Methods:** This study spanned from December 1, 2020 to January 23, 2021 and included the populations of Ontario (including Toronto) and Quebec (including Montreal). The intervention of interest was a nighttime curfew implemented across Quebec on January 9, 2021. Unadjusted and adjusted difference-in-differences models (DID) were used to measure the incremental impact of the curfew on nighttime mobility in Quebec as compared to Ontario.

**Results:** The implementation of the curfew was associated with an immediate reduction in nighttime mobility. The adjusted DID analysis indicated that Quebec experienced a 31% relative reduction in nighttime mobility (95%CI: -36% to -25%) compared to Ontario, and that Montreal experienced a 39% relative reduction compared to Toronto (95%CI: -43, -34).

**Discussion:** However, this natural experiment among two neighbouring provinces provides useful evidence that curfews lead to an immediate and substantial decrease nighttime mobility, particularly in these provinces’ largest urban areas hardest hit by COVID-19.

## Background

The effects of the COVID-19 pandemic have been highly variable across countries due to differing governmental and societal responses.^1^ Among non-pharmaceutical interventions, individual movement restrictions have been among the most impactful for controlling case growth.^2^ Nighttime curfews have been implemented in certain regions (New Orleans, France, French Guyana),^3^ but their incremental effect is understudied. In the second wave of COVID-19, Canada’s two largest and adjacent provinces implemented lockdown restrictions with (Quebec) and without (Ontario) a nighttime curfew, providing a natural experiment to study the association between curfews and mobility. We examine whether curfews lead to decreased nighttime mobility, and whether the impact differed across socio-economic status (SES) groups.

## Methods

This study spans from December 1, 2020 to January 23, 2021 and includes the populations of Ontario (including Toronto) and Quebec (including Montreal). The intervention of interest was a nighttime curfew implemented across Quebec on January 9, 2021. We used anonymized mobility data from Veraset, drawn from location-based services across third-party mobile applications on both Apple and Android devices, after users have consented to use of their anonymized data; the data represents approximately 1% of the population. Mobility data was used to estimate the daily proportion of devices travelling ≥500 metres during curfew (8PM-5AM) and non-curfew (5AM-8PM) hours. Unadjusted and adjusted difference-in-differences (DID) models were used to compare daily mobility values at the provincial (Quebec versus Ontario) and city-level (Montreal versus Toronto). The unadjusted DID models measured the relative change in mobility before and after January 9, 2021, for Ontario/Toronto (reference) versus Quebec/Montreal, while the adjusted models included additional adjustments for day of the week and contemporaneous effects. Canadian Census data were used to group neighbourhoods into tertiles based on median household income and proportion working in essential services. Ethics approval was obtained from the University of Toronto Research Ethics Board through the Modeling Consensus Table, a provincial working group developing evidence for the COVID-19 response that operates with the support of the Ontario Ministry of Health, Ontario Health, and Public Health Ontario.

## Results

In the pre-curfew period, both jurisdictions experienced similar daytime and nighttime mobility trends, followed by a gradual increase in daytime (but not nighttime) mobility over the first week of the year (Figure 1). The implementation of curfew in Quebec on January 9, 2021 was associated with an immediate, steep reduction in nighttime mobility (Figure 1). The adjusted DID analysis indicated that Quebec experienced a 31% relative reduction in nighttime mobility (95%CI: -36% to -25%) and an 8.7% relative increase in daytime mobility (95%CI: 4.9% to 13%) compared to Ontario; results were similar at the city-level (Table 1). In Montreal, the reductions in nighttime mobility varied by tertiles of neighbourhood-level household income (highest: -40.5%, middle: -38.7%, lowest: -33.7%) and essential workers (highest: -32.6%, middle: -40.6%, lowest: -39.3%).

**Table 1.** Proportion of devices moving 500 metres or more between December 1, 2020 and January 23, 2021.

| | Proportion of devices moving $\geq 500\text{m}$ | | | Difference-in-differences (%) | |
| --- | --- | --- | --- | --- | --- |
|  | Pre-curfew<br>(Dec 1 to Jan 8) | Post-curfew<br>(Jan 9 to Jan 23) | Pre-Post<br>Difference | Unadjusted* | Adjusted** |
| Province-level |  |  |  |  |  |
| Nighttime (8PM-5AM) |  |  |  |  |  |
| Ontario | 0.138 | 0.115 | -0.023 | Reference | Reference |
| Quebec | 0.128 | 0.078 | -0.050 | -21 | -31 (-36, -25) |
| Daytime (5AM-8PM) |  |  |  |  |  |
| Ontario | 0.380 | 0.383 | 0.003 | Reference | Reference |
| Quebec | 0.384 | 0.421 | 0.037 | 8.8 | 8.7 (4.9, 13) |
| City-level |  |  |  |  |  |
| Nighttime (8PM-5AM) |  |  |  |  |  |
| Toronto | 0.091 | 0.078 | -0.013 | Reference | Reference |
| Montreal | 0.090 | 0.050 | -0.040 | -30 | -39 (-43, -34) |
| Daytime (5AM-8PM) |  |  |  |  |  |
| Toronto | 0.269 | 0.269 | 0.000 | Reference | Reference |
| Montreal | 0.292 | 0.312 | 0.020 | 6.8 | 7.5 (3.4, 12) |

\* The unadjusted estimate is based on the simple difference in the pre-post differences; \*\* the adjusted estimate is based on a time series model that also adjusted for day of the week and contemporaneous effects<sup>4</sup>

**Figure 1.**
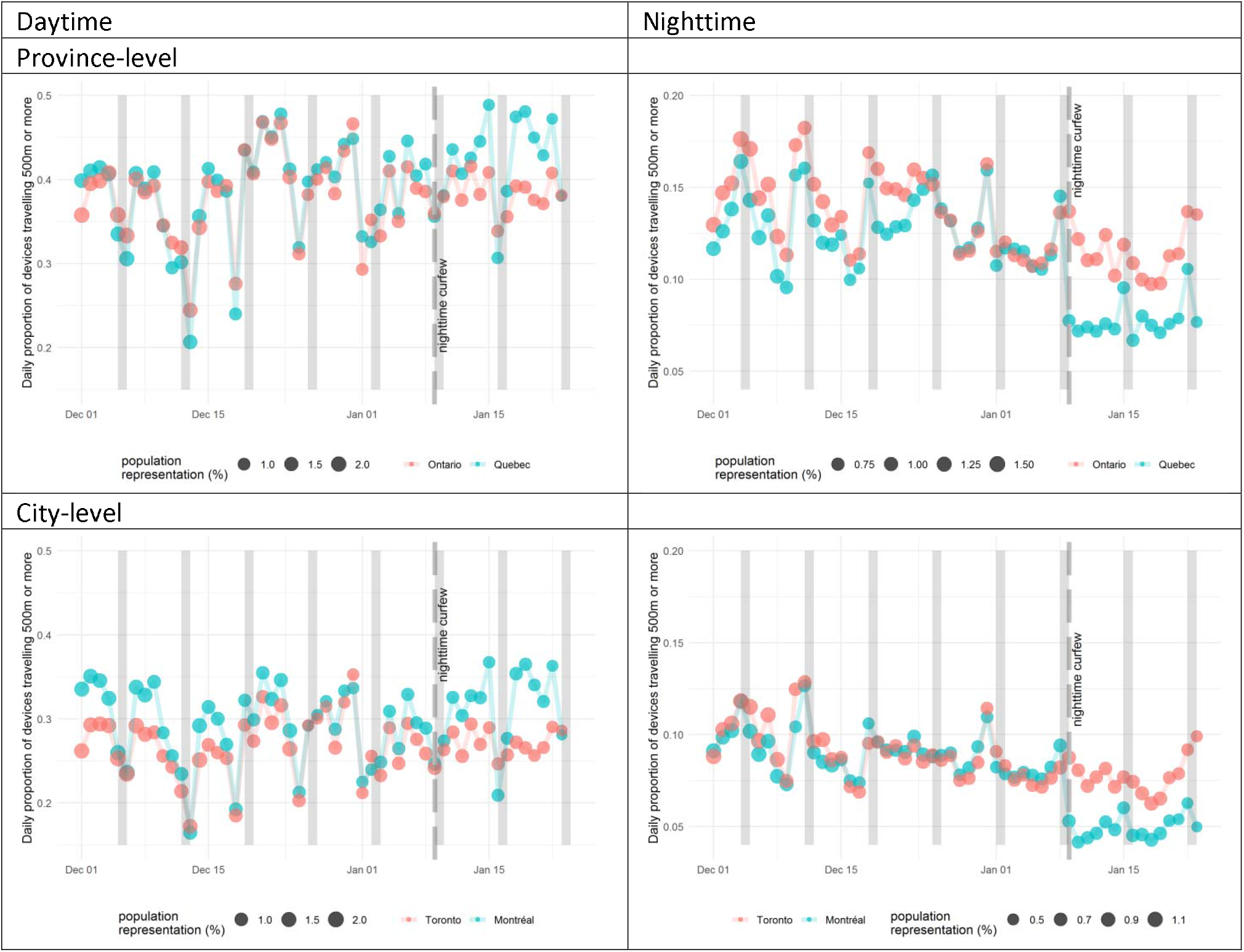
Daily proportion of devices moving 500m or more during the daytime (left panels) and nighttime (right panels) prior to and following implementation of the nighttime curfew in Quebec (including Montreal) [blue dots] on January 9^th^, as compared to absence of curfew in Ontario (including Toronto) [red dots]. Vertical bars show Saturday and Sunday daytime (left panel), and Friday and Saturday nighttime (right panel). Note that schools reopened in Quebec on January 11, 2021 but only reopened provincewide in Ontario on February 16, 2021.

## Discussion

Nighttime curfews led to a 31% relative reduction in nighttime mobility in Quebec (39% in Montreal). The impact appeared somewhat inequitable, with smaller mobility reductions seen in neighbourhoods with lower SES and higher proportions of essential workers that have borne the brunt of COVID-19 incidence and mortality.^6^ Previous studies in France suggest that curfews may be associated with attenuation in COVID-19 growth rates.^7^ Our results suggest that curfews can successfully reduce nighttime mobility. Nighttime curfews may be efficient because they reduce mobility when people are more likely to have social interactions with family members, friends, and acquaintances, while having relatively little impact on work and outdoor exercise.^3^ Study limitations include missing data on movement among those without a GPS or WiFi-enabled device, mobility being only a proxy for social contact, and differing policies affecting daytime mobility (e.g. schools reopened in Quebec on January 11, 2021 but only reopened provincewide in Ontario on February 16, 2021). However, this natural experiment among two neighbouring provinces provides useful evidence that curfews lead to an immediate and substantial decrease nighttime mobility, particularly in these provinces’ largest urban areas hardest hit by COVID-19.

## Data Availability

Aggregated province-level and city-level data used for the analysis are available upon request.

## Notes

### Competing Interest Statement

The authors have declared no competing interest.

### Funding Statement

This study received no external funding.

### Author Declarations

Ethics approval was obtained from the University of Toronto Research Ethics Board through the Modeling Consensus Table, a provincial working group developing evidence for the COVID-19 response that operates with the support of the Ontario Ministry of Health, Ontario Health, and Public Health Ontario.

